# Endoscopically visible Randall’s plaques are an independent predictor of future stone events

**DOI:** 10.1101/2025.09.25.25336682

**Authors:** Natalie Meyer, Jean Lee, Victoria Liu, Lejla Pepic, Robert Pearce, Kevin Li, Kevin Shee, Jorge Mena, Justin Ahn, David Bayne, Marshall Stoller, Tom Chi, Wilson Sui, Heiko Yang

## Abstract

**Introduction and objectives:** Randall’s plaques have been long thought to be precursors for kidney stone formation, but how their presence impacts future stone recurrence is not known. The aim of this study was to determine whether patients with endoscopically visible plaques were more likely to have subsequent stone events compared to those without.

**Methods:** The presence of Randall’s plaque in adult patients was prospectively assessed during endoscopic cases as part of the Registry for Stones of the Kidney and Ureter (ReSKU). In each case, the visible Randall’s plaque burden was scored by the attending surgeon as “none,” “minimal,” or “many.” Data regarding stone composition, 24-hour urine tests, and subsequent stone events on follow up were collected. Stone events were defined as patient-reported stone passage or any primary operation for kidney stones; second-look or staged operations were not counted as separate stone events.

**Results:** We identified 673 subjects with a Randall’s plaque assessment, of which 78 had “none,” 306 had “minimal,” and 289 had “many.” Despite having no significant differences in initial stone burden, subjects with visible Randall’s plaque (“minimal” or “many”) had a higher relative risk of stone recurrences after surgery compared to those without (RRR 1.711 (1.121-2.611), p = 0.01). Interestingly, no significant differences in 24-hour urine analytes were observed. Subjects with visible plaque were more likely to have calcium-based stones (84% vs 58%, p < 0.001).

**Conclusion:** The presence of endoscopically visible Randall’s plaque is associated with an increased risk of stone recurrence independent of urine composition.

## INTRODUCTION

Nephrolithiasis is an increasingly prevalent condition, affecting approximately 10.9% of men and 9.4% of women in the United States.^1^ Preventing subsequent stone recurrence, which affects around 17% of all new stone formers and increases up to 60% after multiple stone events, is a major clinical challenge.^2, 3^ While metabolic and dietary interventions are commonly recommended for reducing kidney stone recurrence, their long-term effectiveness remains limited.^4^ There is an important need to better understand the mechanistic factors that contribute to stone formation and recurrence.

The majority of first-time stone formers produce calcium-based calculi, with calcium oxalate (CaOx) being the predominant composition in approximately 76% of cases.^5^ In the 1930s, Alexander Randall proposed that calcium oxalate stones often originate on subepithelial papillary plaques composed of interstitial hydroxyapatite, now known as Randall’s plaques.^6,7^ Contemporary studies have validated this mechanism, identifying Randall’s plaques as common nucleation sites for CaOx stones.^8^ Histologic analysis suggests that plaque formation begins in the basement membranes of the thin limbs of Henle’s loop and extends into the papillary interstitium.^9^ Taken together, there is a growing interest in the visualization of plaque burden as a potential biomarker for long-term health implications.^10, 11^

In this study, we aimed to evaluate the association between endoscopically visualized Randall’s plaques and kidney stone recurrence using clinical and metabolic data. We hypothesized that the presence of Randall’s plaques at the time of surgery would be significantly associated with calcium stone recurrence, thereby offering a potential intraoperative indicator for long-term patient risk stratification and targeted prevention.

## METHODS

### Study cohort

Prospectively collected data from adult subjects (aged ≥ 18 years) enrolled in the Registry for Stones of the Kidney and Ureter at the University of California, San Francisco were queried. All participants provided written consent, and the study was approved by the Institutional Review Board and Committee on Human Research (Protocol 14-14533).

Subjects who had undergone endoscopic stone surgery (ureteroscopy and/or percutaneous nephrolithotomy) from 2015 to 2023 were included in the analysis. Demographics, comorbidity data, and disease-specific evaluation (stone size, number, and location) were collected in the initial visit.

### Measured variables and outcomes

During the initial operation, a visual assessment of Randall’s plaque burden was performed and immediately documented in a standardized Brief Operative Note by the attending surgeon. Randall’s plaque burden was subjectively scored as “none”, “minimal,” or “many.” Surgically removed stone fragments were sent to the University of California, San Francisco clinical laboratory where stone analysis was conducted using a standard method based on infrared spectroscopy.

Twenty-four hour urine analytes were abstracted and all studies were performed by a specialized laboratory (Litholink Corporation, Chicago, IL). Twenty-four hour urine volume, calcium, oxalate, citrate, pH, phosphorous, chloride, sodium, potassium, magnesium, sulfate, urea nitrogen, uric acid, creatinine and creatinine per kilogram were abstracted. The supersaturation index of calcium oxalate (SSCaOx), calcium phosphate (SSCaPhos), and uric acid (SSUA) were reported by Litholink™. Only the results of the patient’s initial 24-hour urine were included in this study. Inadequate collections were defined by mg creatinine excretion per kg patient weight (<11.9 or >24.4 for males and < 8.7 mg and >20.3 for females). Patients with incomplete clinical or follow-up data were excluded in addition to patients with cystinuria or who had less than 180 days of follow up.

The primary outcome was stone recurrence. Stone recurrence was defined as a subsequent stone event >90 days after initial presentation by patient report of stone passage or procedural intervention. This included procedural interventions reported by patients that occurred outside of our institution. Planned second-look or staged procedures were excluded. Follow up time was defined by the time between their stone surgery to their final follow up clinic visit. Stone event rate was calculated by the number of stone events divided by their total follow up time.

### Statistical analysis

Analysis of variance (ANOVA) was used for continuous variables and chi-square analysis for categorical variables. Multivariable analyses were performed first using poisson regression to obtain adjusted relative risk ratios. Follow up time was used as an offset variable and the model was adjusted for age, race and gender in addition to history of coronary artery disease, hyperlipidemia, type 2 diabetes and presence of Randall’s plaque. These results were compared to negative binomial regressions and due to the improved log likelihood ratio, to minimize dispersion, the negative binomial regression was used as the final model. All statistics were performed using SPSS™ v27.

## RESULTS

Of the 673 patients included in the study cohort, 78 had no visible plaque (“none”), 306 had “minimal”, and 289 had “many” visible Randall’s plaques (Table 1). We did not detect any significant differences in age, sex, race, or comorbidity among these groups.

**Table 1.**
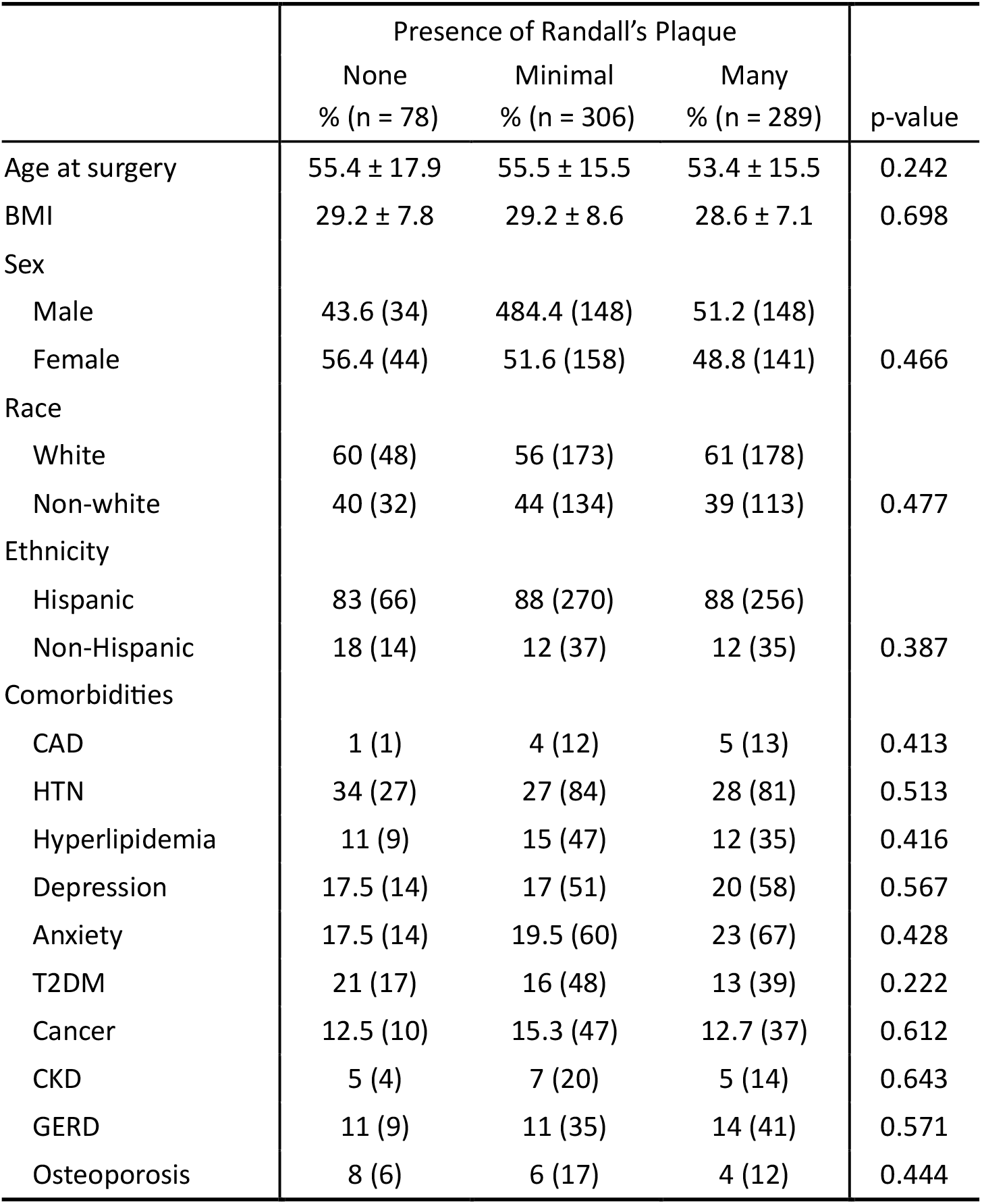
Demographic characteristics of the study cohort.

|  | Presence of Randall's Plaque |  |  | p-value |
| --- | --- | --- | --- | --- |
|  | None<br>% (n = 78) | Minimal<br>% (n = 306) | Many<br>% (n = 289) |  |
| Age at surgery | 55.4 ± 17.9 | 55.5 ± 15.5 | 53.4 ± 15.5 | 0.242 |
| BMI | 29.2 ± 7.8 | 29.2 ± 8.6 | 28.6 ± 7.1 | 0.698 |
| Sex |  |  |  |  |
| Male | 43.6 (34) | 484.4 (148) | 51.2 (148) | 0.466 |
| Female | 56.4 (44) | 51.6 (158) | 48.8 (141) |  |
| Race |  |  |  |  |
| White | 60 (48) | 56 (173) | 61 (178) | 0.477 |
| Non-white | 40 (32) | 44 (134) | 39 (113) |  |
| Ethnicity |  |  |  |  |
| Hispanic | 83 (66) | 88 (270) | 88 (256) | 0.387 |
| Non-Hispanic | 18 (14) | 12 (37) | 12 (35) |  |
| Comorbidities |  |  |  |  |
| CAD | 1 (1) | 4 (12) | 5 (13) | 0.413 |
| HTN | 34 (27) | 27 (84) | 28 (81) | 0.513 |
| Hyperlipidemia | 11 (9) | 15 (47) | 12 (35) | 0.416 |
| Depression | 17.5 (14) | 17 (51) | 20 (58) | 0.567 |
| Anxiety | 17.5 (14) | 19.5 (60) | 23 (67) | 0.428 |
| T2DM | 21 (17) | 16 (48) | 13 (39) | 0.222 |
| Cancer | 12.5 (10) | 15.3 (47) | 12.7 (37) | 0.612 |
| CKD | 5 (4) | 7 (20) | 5 (14) | 0.643 |
| GERD | 11 (9) | 11 (35) | 14 (41) | 0.571 |
| Osteoporosis | 8 (6) | 6 (17) | 4 (12) | 0.444 |

Analysis of stone disease characteristics revealed no significant differences in cumulative stone size, laterality, or number of stones among the groups (Table 2), although subjects with “many” Randall’s plaques more frequently underwent ureteroscopy alone than PCNL or a combination surgery (p < 0.001).

**Table 2.** Disease characteristics of the study cohort.

|  | Presence of Randall's Plaque |  |  | p-value |
| --- | --- | --- | --- | --- |
|  | None<br>% (n = 78) | Minimal<br>% (n = 306) | Many<br>% (n = 289) |  |
| Mean stone burden<br>(mm) | 28 ± 28 | 27 ± 27 | 26 ± 26 | 0.868 |
| Laterality |  |  |  |  |
| Unilateral | 30 (24) | 28 (86) | 30 (87) | 0.862 |
| Bilateral | 70 (56) | 72 (221) | 70 (204) |  |
| Number of stones |  |  |  |  |
| None | 3 (2) | 3 (7) | 3 (7) | 0.194 |
| Solitary | 35 (20) | 38 (87) | 28 (59) |  |
| 1 to 3 | 26 (15) | 33 (74) | 31 (66) |  |
| > 3 | 36 (21) | 26 (60) | 37 (79) |  |
| Surgery type |  |  |  |  |
| URS | 46 (37) | 46 (142) | 67 (196) | <0.001 |
| PCNL | 33 (26) | 39 (117) | 18 (53) |  |
| URS + PCNL | 21 (17) | 16 (48) | 14 (42) |  |
| Stone composition | % (n = 33) | % (n = 161) | % (n = 149) |  |
| Other | 42 (14) | 17 (28) | 15 (22) | <0.001 |
| Calcium | 58 (19) | 83 (133) | 85 (128) |  |

Subjects with visible Randall’s plaques (“minimal” or “many”) were more likely to have calcium-based stones on stone analysis compared to those with no Randall’s plaques (83% and 85% vs 58%, respectively, p = 0.021) (58%). Interestingly, no significant differences in 24-hour urine composition were observed among the groups, except for urinary creatinine, which was significantly higher in the “none” group (p = 0.021) (Table 3). Urinary calcium did trend slightly higher with increasing plaque burden (“none”: 186.3 ± 107.5, “minimal”: 194.7 ± 117, “many”: 210.4 ± 128.7 but was not statistically significant (p = 0.446)

**Table 3.**
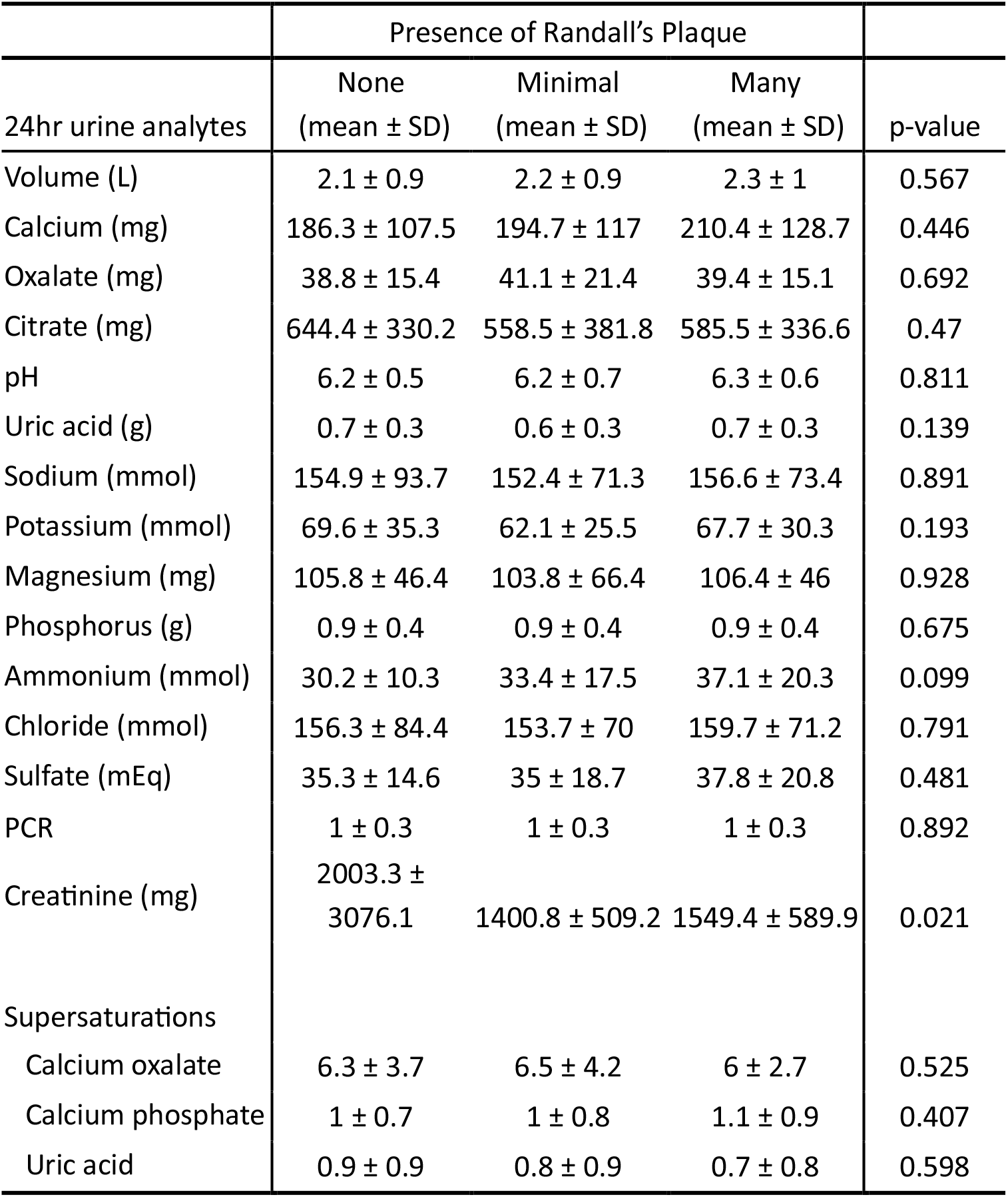
Comparison of 24-hour urine parameters in the study cohort.

Over a five-year follow-up period, patients with visible (“minimal” or “many”) Randall’s plaques experienced a greater number of stone events (Figure 1). In a negative binomial regression, the presence of visible Randall’s Plaque was an independent predictor of stone recurrence with a relative risk ratio of 1.7 (1.2-2.6) (p = 0.013) (Table 4).

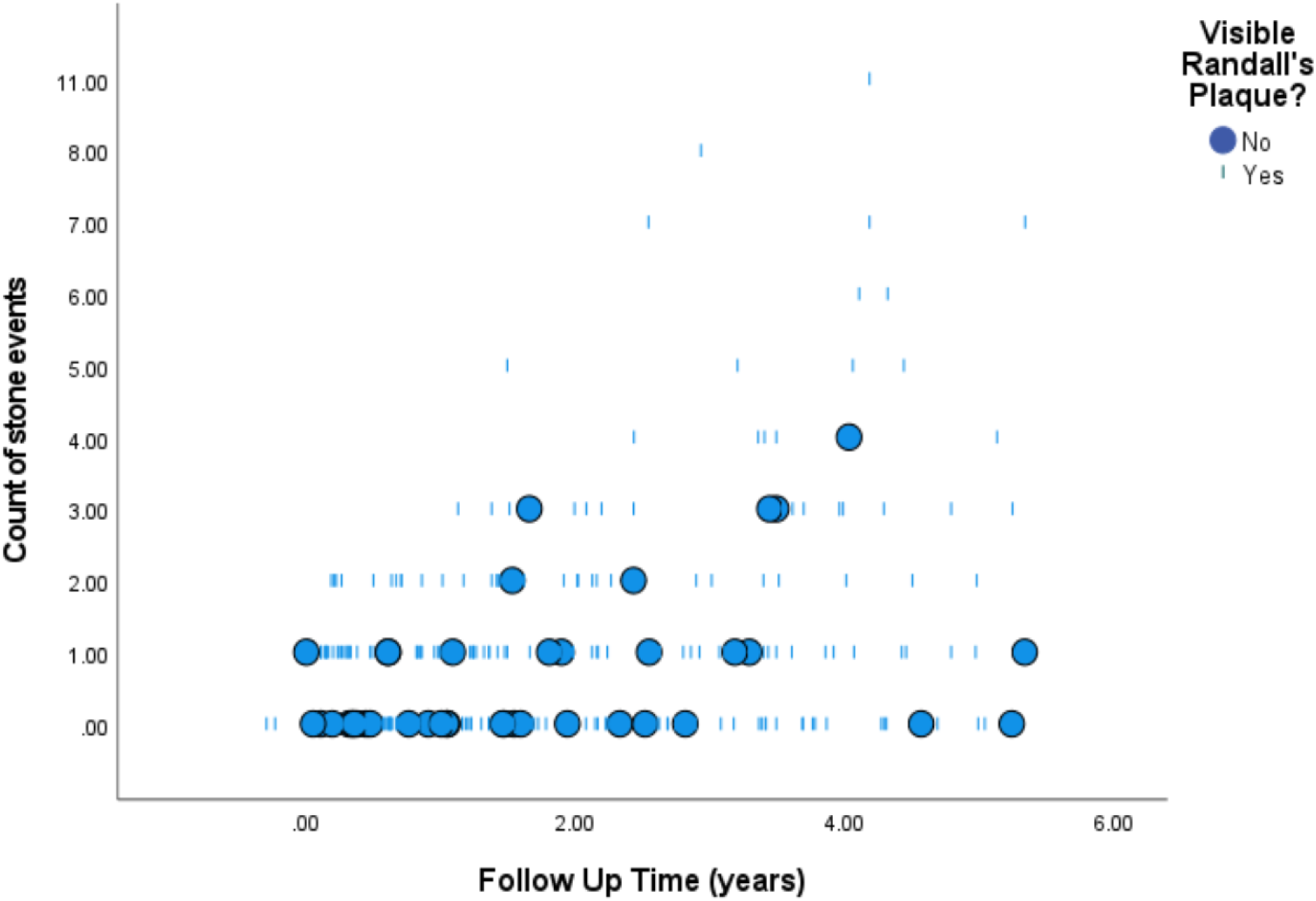

**Table 4.** Multivariable model of stone recurrence.

|  | RRR (95%CI) | p-value |
| --- | --- | --- |
| Age at surgery | 0.983 (0.976 - 0.991) | <0.001 |
| BMI | 1.009 (0.992 - 1.025) | 0.298 |
| Sex |  |  |
| Male | ref | Ref |
| Female | 1.011 (0.796 - 1.283) | 0.929 |
| Race |  |  |
| White | ref | ref |
| Non-white | 0.979 (0.769 - 1.245) | 0.86 |
| Comorbidities |  |  |
| CAD | 1.383 (0.716 - 2.67) | 0.334 |
| Hyperlipidemia | 0.533 (0.384 - 0.739) | <0.001 |
| T2DM | 2.095 (1.444 - 3.038) | <0.001 |
| Randalls plaque |  |  |
| None | ref | ref |
| Visible | 1.711 (1.121 - 2.611) | 0.013 |

## DISCUSSION

Stone recurrence remains a key challenge in the management of urolithiasis. Recurrence rates rise significantly with subsequent episodes, beginning at 17% after the first symptomatic episode and reaching 60% after the fourth or higher episode.^2,3^ As stone diseases prevalence grows, the identification of patients at high risk of recurrence is crucial to enable timely and targeted prevention strategies. Although several predictive models for recurrence have been proposed, their clinical utility remains limited. For example, the Recurrence of Kidney Stone (ROKS) nomogram yields only modest predictive performance, with a concordance index (C-index) of 0.681.^2, 12^ Moreover, many of these models do not offer mechanistic insight for stone prevention.

Real-time visualization of the renal papillae and assessment of Randall’s plaque during endoscopic procedures could help bridge the scientific gap in understanding and serve as a simple clinical tool for identifying patients who are at risk for future stone events. Our findings demonstrate that patients with visible Randall’s Plaque experienced more frequent stone events over the five-year follow up period independent of 24-hour urine composition, comorbidities, and demographics. Among those with more than four stone events on follow-up, all exhibited the Randall’s Plaque phenotype, further supporting the hypothesis that Randall’s Plaque serves as a persistent nidus for future stone formation.

Given that Randall’s Plaque presence cannot be reliably predicted based on baseline demographic or clinical characteristics and cannot be reliably seen on non-invasive imaging, intraoperative endoscopic assessment remains essential for phenotype evaluation. Systematic evaluation and documentation of the Randall’s Plaque phenotype during surgery offers an opportunity to tailor long-term management strategies aimed at reducing recurrence risk in these patients. Future research should focus on elucidating the molecular and cellular mechanisms underlying stone formation in patients with Randall’s Plaque, which may inform the development of phenotype-specific preventative or therapeutic interventions.

This study had several limitations that should be considered when interpreting the findings. First, it was conducted at a single institution, with data collected from cases performed by four different surgeons. Although attending physicians assessed Randall’s Plaque burden, the subjective classification of “minimal” versus “many” may have significant variability. To address this, we combined these categories for the analysis presented in Figure 1 and the recurrence rates in Table 4. Additionally, we were unable to control for true stone-free rate after surgery, raising the possibility that residual stone fragments may have contributed to subsequent stone events. Follow-up data may also be subject to bias, as some patients were lost to follow-up due to changes in care or lack of documented recurrence. Finally, the small sample size limited our ability to perform Kaplan-Meier analysis to graphically represent recurrence probability over time.

## CONCLUSION

The presence of endoscopically visible Randall’s Plaque on the renal papillae is independently and significantly associated with an increased risk of kidney stone recurrence. Identification of this phenotype during surgery may offer a valuable tool for stratifying patient risk and guiding long-term management strategies.

## Data Availability

All data produced in the present study are available upon reasonable request to the authors

## AUTHOR CONTRIBUTIONS

NM, WS, and HY drafted the manuscript.

NM, JL, VL, LP, RP, KL, KS, and JM collected the data.

WS and HY analyzed the data and created the figures and tables.

JA, DB, MS, TC, WS, and HY conceptualized the project.

All authors reviewed the manuscript.

